# An outbreak of SARS-CoV-2 among residents of a senior housing apartment building in Massachusetts

**DOI:** 10.1101/2022.06.12.22276282

**Authors:** Victoria M. Selser, Lauren E. Saunders, Catherine M. Hoover, Kwonjune J. Seung

## Abstract

Independent senior housing is a high risk setting for COVID-19 and should be prioritized for outbreak investigation and response. During a COVID-19 outbreak in Massachusetts from December 2021 to January 2022, 47% of residents in a 31-unit independent senior housing apartment building in Massachusetts were infected. Residents that had received a booster dose of an mRNA COVID-19 vaccine had a lower attack rate than those who had received two doses. Comprehensive response campaigns that include education, testing, treatment referrals, and vaccination can be effective at reducing morbidity and mortality and preventing future outbreaks.

## Background

Since the beginning of the pandemic, congregate care facilities for the elderly have been known to be epidemiological hot spots for COVID-19, but there is comparatively little known about independent senior housing. In the United States, congregate care facilities for the elderly include nursing homes, long term care, assisted living, and memory care. Whereas congregate care facilities provide residents with varying degrees of medical and supportive care, the primary focus of independent senior housing is the social needs of residents. Independent senior housing may be advertised as “retirement communities”, “over 55”, or “independent living”. Congregate care facilities have had frequent and deadly outbreaks of COVID-19, and like other health care facilities, often have strict policies aimed at preventing SARS-CoV-2 transmission (e.g. vaccine mandates, regular screening, restricted visitors). This is not the case for independent senior housing, where COVID-19 related policies are similar to other apartment buildings.

The Commonwealth of Massachusetts, located in the northeast of the United States of America, has a population of close to seven million people. The Montachusett Public Health Network (MPHN) is a regional public health collaborative of 13 cities and towns in Worcester County, Massachusetts, with a combined population of approximately 175,000. In December 2021, a 31-unit independent senior apartment building experienced an outbreak of COVID-19 that infected 47% of the residents. In this report, we present the epidemiology of the outbreak and the public health response.

## Methods

### Case investigation

A confirmed case of COVID-19 was defined as a person with a positive SARS-CoV-2 nucleic acid amplification test result or positive rapid antigen test result during the period of the outbreak. A presumptive case of COVID-19 was defined as a person who reported symptoms consistent with COVID-19 during the period of the outbreak, but who did not seek testing. Laboratories electronically report SARS-CoV-2 test results to the Massachusetts Virtual Epidemiologic Network (MAVEN), the statewide disease surveillance and case management system (1). As part of the investigation and response, MPHN interviewed residents of the apartment building to collect information about demographics, symptoms, vaccination, and exposure source. Additional information about vaccination status was obtained from the Massachusetts Immunization Information System (MIIS) (2).

## Data analysis

Attack rates were compared by demographic characteristics, vaccination status, booster status, vaccine product, and time since vaccination. Differences between groups were assessed using chi-square or Fisher’s exact tests using SAS® (SAS Institute). P-values were calculated with the chi-square test (when all cell sizes ≥5) or Fisher’s exact test (when any cell size <5).

## Ethics

COVID-19 is a reportable disease in Massachusetts and case investigation and data collection were conducted for public health purposes. Data were stripped of direct identifiers prior to analysis. The Mass General Brigham Institutional Review Board determined that this study was secondary research for which consent was not required.

## Results

The MPHN became aware of this outbreak after noticing multiple laboratory confirmed cases with the same address in MAVEN and reached out to building management to coordinate the investigation and response. The apartment building, constructed in 1920, has 31 units across five floors. The first floor has three units and the remaining floors have seven units connected by a single hallway and elevator. Common spaces are on the first floor and include a community room, laundry room, and mail area. Occupancy is restricted to elderly or disabled households and most apartment units are occupied by a single person. There is no central ventilation system linking apartment units.

From December 21, 2021 to January 9, 2022, there were 15 confirmed cases with a positive nucleic acid amplification test result (11) or positive rapid antigen test result (4) (Figure 1). One positive sample was sequenced and was found to be Delta variant AY.47; others were not able to be retrieved for sequencing. Three confirmed cases were hospitalized due to COVID-19, and one case died. There were three presumptive cases who were symptomatic but did not get tested. There was also one confirmed case among the staff, but this person did not reside in the region so full information about the case was not available to the MPHN and is excluded from the analysis. Confirmed and presumptive cases totaled 18, which was 47% of the residents. There were 15 apartment units (48%) with at least one case. Of note, no COVID-19 cases had been reported at this address in the 11 months prior to the discovery of this outbreak.

**Figure 1:**
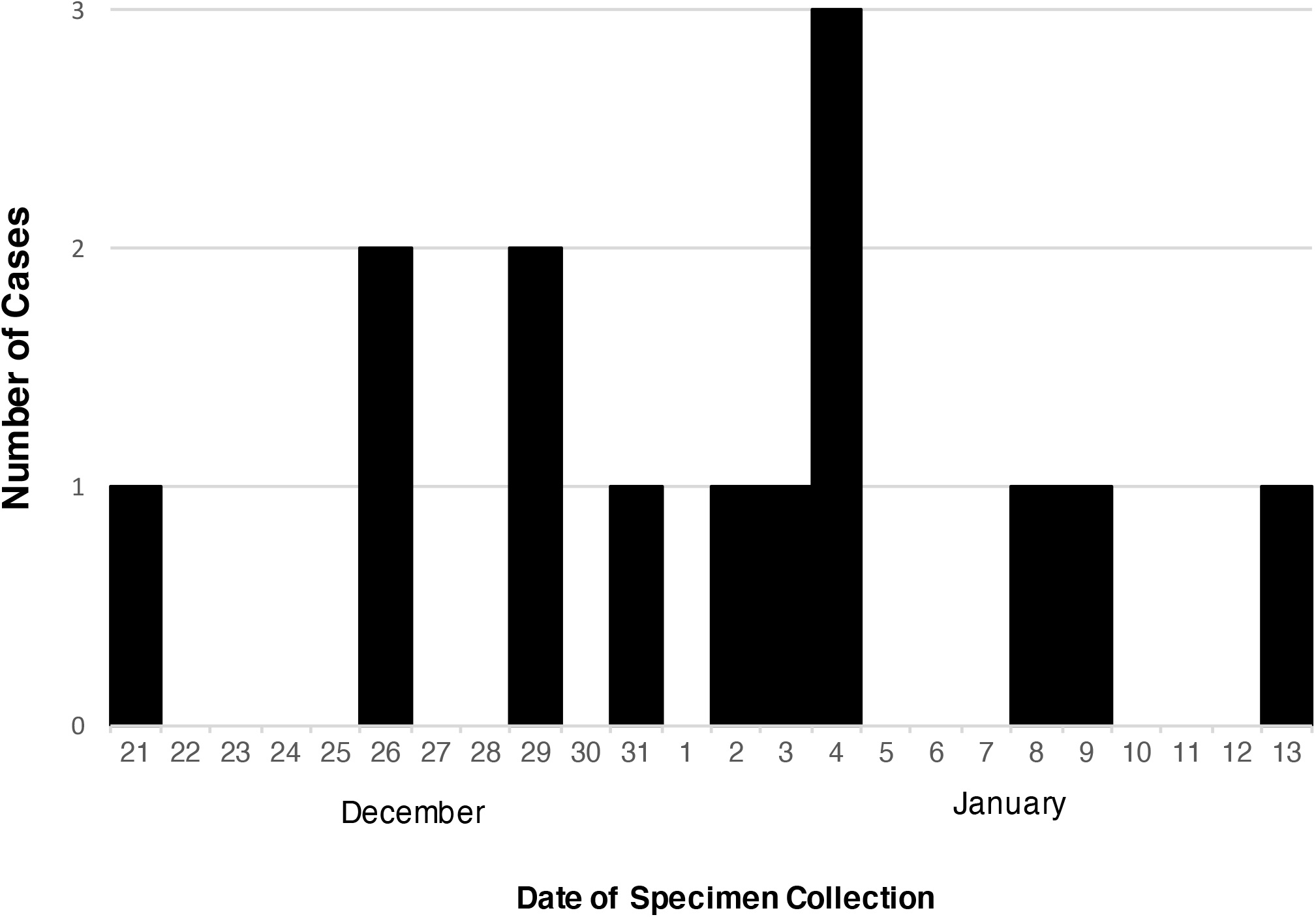
Date of specimen collection for cases positive for SARS-CoV-2 living in an independent senior housing apartment building (N=14*)—Worcester County, Massachusetts, December 2021-January 2022. * Excludes three presumptive cases (defined as a resident who had symptoms consistent with COVID-19 during the period of the outbreak but who did not get tested) and one confirmed case with a missing date of specimen collection.

Interviews suggested that transmission was likely driven by social interactions, particularly in the community room where residents frequently gathered for group activities such as bingo. The community room was closed for two weeks following identification of the outbreak. Transmission pathways between apartment units, such as exhaust vents or drainage pipes, could not be ruled out (3,4).

Of the 38 residents, 32 (84%) had received at least two doses of either the Moderna or Pfizer-BioNTech COVID-19 vaccine a mean of 235 days (range 133 to 294 days) before December 21, 2021, the date of the first positive SARS-CoV-2 test reported in this outbreak. 18 (47%) had received a third dose a mean of 17 days (range -17 to 49 days) before the start of the outbreak. Residents who had received a third dose had a significantly lower attack rate than those who had only received two doses (p = 0.04) (Table 1). There were no infections among the 10 residents who received a third dose more than two weeks before the start of the outbreak (p = 0.003).

**Table 1:** SARS-CoV-2 attack rate among residents of a senior apartment building by vaccination status, product and timing—Worcester County, Massachusetts, December 2021-January 2022.

| Characteristic | Total | Number of cases | Attack rate (%) | P-value* |
| --- | --- | --- | --- | --- |
| **Vaccination status** |  |  |  |  |
| Unvaccinated | 6 | 4 | 66.7% | 1.0 |
| Two doses | 14 | 9 | 64.3% | Ref |
| Three doses | 18 | 5 | 27.8% | 0.04 |
| **Vaccine product** |  |  |  |  |
| Moderna | 17 | 7 | 41.2% | Ref |
| Pfizer-BioNTech | 15 | 7 | 46.7% | 0.75 |
| **Time from third dose to outbreak†** |  |  |  |  |
| < 14 days | 7 | 5 | 71.4% | Ref |
| ≥ 14 days | 10 | 0 | 0% | 0.003 |
| **Sex** |  |  |  |  |
| Female | 23 | 11 | 47.8% | Ref |
| Male | 15 | 7 | 46.7% | 0.94 |
| **Age group** |  |  |  |  |
| ≤ 65 years | 18 | 8 | 44.4% | Ref |
| > 65 years | 20 | 10 | 50% | 0.73 |

\* P-values calculated with chi-square test (when all cell sizes ≥5) or Fisher's exact test (when any cell size <5).
† Date of third dose is missing for one person.

## Discussion

This is the first report of a COVID-19 outbreak in independent senior housing in the medical literature even though such outbreaks are likely to be quite common. Age is the most important risk factor for COVID-19 morbidity and mortality; older adults often suffer from chronic illnesses that are independent risk factors for hospitalization and death from COVID-19 (5). Due to advanced age and comorbidities, COVID-19 outbreaks in independent senior housing are likely to cause high rates of hospitalization and death.

As social distancing restrictions are lifted, increasing social events organized within these settings may trigger cluster transmission. While long-term care and assisted living facilities have strict SARS-CoV-2 infection and prevention control measures in place at the state and federal levels, there are few such policies for independent senior housing despite similar risk profiles of residents. The lack of these measures can result in unchecked spread followed by preventable negative health outcomes.

The severity of outbreaks in independent senior housing can be exacerbated by gaps in vaccination and booster coverage. While 66% of people in the United States have received a primary series of vaccination, less than half have received a first booster dose (6). In this outbreak, residents with a third dose of an mRNA vaccine had a lower attack rate compared to residents with two doses, consistent with data from multiple countries showing the protective effect of a third dose (7,8). Full protection of a booster dose, however, may not be attained until two weeks after its administration.

Independent senior housing should be prioritized for vaccine outreach and clinics; residents are not only high risk but also often experience barriers to access. Physical disability or lack of transportation can limit access to vaccination for many residents. Prior to the vaccine clinics organized by MPHN following this outbreak, there had not been an on-site vaccination opportunity for residents of this apartment building. In addition to the convenience of the on-site vaccine clinics, much of the success of the vaccination effort was due to the use of multiple complementary communication methods such as notices from building management, phone calls, and door-to-door outreach.

To facilitate early detection of outbreaks in this setting, local health departments should proactively monitor the addresses of COVID-19 cases in their jurisdiction for residents of independent senior housing properties. When an outbreak is detected, a comprehensive public health response—including outreach to residents, distribution of masks and tests, referral for treatment and vaccination—will mitigate the severity of the outbreak and help to prevent future ones (Box 1).

In this COVID-19 outbreak in an independent senior housing apartment building in Massachusetts, there was a high rate of severe illness among infected residents and a significantly higher attack rate among unboosted residents. Local health departments should prioritize this setting for proactive vaccine outreach and COVID-19 outbreak investigation and response. Comprehensive response campaigns that include education, testing, treatment referral, and vaccination can be effective at reducing morbidity and mortality and preventing future outbreaks.

## Data Availability

The data produced in the present work is not available to the public.

## Acknowledgements

Community Health Connections Mobile COVID-19 Vaccination Team; Carole Appleton, Jaime Carrillo, Cynthia Chin, Eileen Hackney, Caitlin Kohl, Dale Kohl, Irene Luperon, Anna Wislocki, Montachusett Public Health Network COVID-19 Response Team.

## Conflict of interest and funding

The authors do not have any commercial or other association that might pose a conflict of interest. This study did not receive any funding.

### Box 1

**Public health response to an COVID-19 outbreak in an independent senior housing apartment —Worcester County, Massachusetts, December 2021-January 2022**

- **Door-to-door outreach**. The MPHN COVID-19 Response Team knocked on every door of the apartment building and talked with residents of 21 units (68%). Building management informed residents prior to the outreach with printed notices that were left on each apartment door.
- **Test and mask distribution**. Each resident was provided with KN95 masks, a COVID-19 rapid antigen self-test kit, and educated on how and when to use them.
- **Education**. Each apartment was provided with an educational packet with information on COVID-19 booster eligibility and monoclonal antibody therapy.
- **Vaccination**. Two vaccine clinics were held in the apartment building community room following the outbreak. Building management left a notice on each apartment door notifying residents of the clinics. Ten residents and two staff members were vaccinated at the clinics.
- **Phone outreach**. Building management provided contact information for all heads of households, and units who were not reached during door-to-door outreach were called.
- **Treatment referrals**. Cases were educated about monoclonal antibody treatment and encouraged to contact their primary care provider if they met eligibility criteria. Residents who did not have or were unable to reach a provider were connected with a local monoclonal antibody treatment center. As a result of this process, two residents received monoclonal antibody treatment.

## Notes

### Competing Interest Statement

The authors have declared no competing interest.

### Author Declarations

The Mass General Brigham Institutional Review Board waived ethical approval for this work.

